# Feasibility & Acceptability of a co-created pragmatic walkability program to promote mobility in older adults with fear of falling: *A question of season and mobility profile?*

**DOI:** 10.64898/2026.09.03.26361963

**Authors:** Marianne Laliberté, Hanane Benabdallah, Kim Lestage, Marie-Soleil Cloutier, François Routhier, Martin Lavallière, Mylène Aubertin-Leheudre

**Affiliations:** Université du Québec à Montréal (UQAM), Faculté des Sciences, Département des Sciences de l’Activité Physique, Groupe de Recherche en Activité Physique Adaptée (GRAPA), Montréal, Canada; Centre de Recherche de l’Institut Universitaire de Gériatrie de Montréal (CRIUGM), Montréal, Canada; Santé-Quebec Montérégie Est, Direction de la Santé Publique, Longueuil, Canada; Institut National de la Recherche Scientifique (INRS), Centre Urbanisation Culture Société, Montréal, Canada; Université Laval, École des sciences de la réadaptation, Québec, Canada; Centre interdisciplinaire de recherche en réadaptation et réintégration sociale, Santé Québec Capitale-Nationale – Universitaire, Québec, Canada; Université du Québec à Chicoutimi (UQAC), Département des sciences de la santé, Lab BioNR et CISD, Chicoutimi, Canada

**Author notes:** **Corresponding author:** Aubertin-Leheudre Mylène, PhD, UQAM, Faculté des Sciences, Département des Sciences de l’Activité Physique 141 av Président Kennedy (#SB 4615) H3C3P8 Montréal Québec-Canada.

**Keywords:** exercise intervention, implementation, real-life setting, aging, community-dwelling population

## Abstract

**Background:** Fear of falling may limit mobility in older adults, particularly during winter. Pragmatic, community-based interventions are needed to support safe, sustainable walking.

**Objective:** To assess the feasibility and acceptability of a co-created intervention (MABIDA) promoting utilitarian and recreational walking among older adults.

**Method:** Fifty-two older adults with fear of falling participated in a 12-week group intervention (summer/winter) consisting of one 60-minute supervised walking session/week, incorporating exercises. Walking group level was assessed using a subjective decisional tree.

**Results:** Forty-one participants (73.3±5.7years) completed the study (dropout: winter only). Retention and adherence exceeded 75%. Intervention settings were considered appropriate (≥81%). The decisional tree correctly classified 72% of participants. Participants reported satisfaction (82%) and enjoyment (81%). No differences were observed between seasons or walking groups (p>0.05). Group walking was the main facilitator (59%), while weather was the main barrier (17%).

**Conclusion:** MABIDA appears feasible, acceptable, and adapted across seasons and walking speed profiles.

## INTRODUCTION

Mobility is a fundamental component of daily life and becomes an increasing concern with aging (Webber et al., 2010). Indeed, a restriction of mobility has been linked to adverse health outcomes, including poorer physical performance (Kuspinar et al., 2020), increased risk of falls (Gonçalves et al., 2024), poorer physical health and quality of life (Rantakokko et al., 2016), higher rates of hospitalization (Johnson et al., 2020) and increased mortality (Kennedy et al., 2017), as it reflects social participation (Kuspinar et al., 2020) and engagement in leisure and physical activity (Peel et al., 2005).

At the societal level, mobility is a central determinant of *aging in place*, enabling older adults to remain safely and independently in their own homes and communities for as long as possible (Bigonnesse & Chaudhury, 2020). Given the demographic shift toward an aging population, promoting and maintaining mobility has become a public health priority to reduce healthcare utilization and support social participation among older adults.

Walking is a fundamental component of mobility. It serves both utilitarian purposes (e.g. going to grocery stores, libraries, shops, etc.) and recreational purposes (e.g. leisure, social walking; (Saelens & Handy, 2008)). Moreover, walking is considered a physical activity (light to vigorous according to the intensity) and is known to induce better functional capacity (Albert et al., 2015), and is associated with better physical health (Shumway-Cook et al., 2005) and greater social engagement/participation (Nemoto et al., 2021). Despite these recognized benefits, walking behaviour and walking life space area tend to decline with advancing age, contributing to a progressive loss in physical performance such as reduced strength, balance and endurance. Musculoskeletal pain, health issues and cognitive decline are some of the well-documented contributors to this reduction.

Falls (Rantakokko et al., 2013) and fear of falling (Auais et al., 2017) are also major determinants of mobility reduction in older populations. Environmental conditions can further exacerbate this phenomenon. Adverse weather, particularly winter conditions characterized by snow, ice and reduced daylight, has been shown to increase fear of falling (Schmidt et al., 2016) and limit outdoor mobility (Clarke et al., 2017) and walking (Bergen et al., 2023; Clarke et al., 2017) among the elderly. These seasonal constraints disproportionately affect older adults and may accelerate mobility decline if not adequately addressed. The urban environment can also be a deterrent to walking in the elderly population, which needs closely spaced resting places like benches and a secure walking environment like sidewalks and safe crossings (Cerin et al., 2017). Moreover, community support to promote elderly walking mobility is often scarce. Interventions have often been delivered in controlled or indoor environments as well as laboratory-based (Nicklas et al., 2020). However, many community-based organizations face persistent constraints in terms of infrastructure and access to expertise, limiting capacity to implement and sustain evidence-based mobility programs (Sims-Gould et al., 2020). Moreover, almost no study has addressed the feasibility of outdoor interventions through different extreme seasons (i.e. icy and snowy winter as well as humid and hot summer). Consequently, there is a pressing need for interventions that are not only evidence-based, but also acceptable, feasible, and scalable within community settings to ensure long-term support for maintaining mobility among older adults. Given growing evidence that co-creation approaches enhance the relevance, acceptability and implementation of community-based programs, such an approach should be prioritized when developing mobility programs for older adults (Slattery et al., 2020).

To address these needs, stakeholders and community-dwelling older adults were involved through walks and interviews to provide their specific needs and barriers (phase I - ORBIT model (Czajkowski et al., 2015)). Following this first step and according to evidence gathered in prior steps (Wheeler-Noiseux & Cloutier, 2026), a pragmatic walking group program through a community organization called MABIDA, which aimed to improve the mobility of older adults with fear of falling, was co-created (phase I - ORBIT model (Czajkowski et al., 2015)).

Thus, this study aimed to assess the feasibility and acceptability of the MABIDA program throughout Canadian seasons in older adults with fear of falling. Based on the literature, we hypothesized that the co-created MABIDA program would be feasible and acceptable for older adults with fear of falling. More specifically, the feasibility and acceptability should be higher during the summer season (fewer environmental/external barriers; (Bergen et al., 2023; Clarke et al., 2017)) and in older adults with slow walking speed (higher need; (Delaire et al., 2021)).

## METHOD

### Design

Action-oriented phase II research (ORBIT model) was conducted in the city of Longueuil (Quebec, Canada). All procedures were approved by the institutional human research ethics committee [*Comité d’éthique en recherche avec des êtres humains (CER)* at the *Institut national de la recherche scientifique (INRS)]* and were conducted in accordance with the principles in the *Declaration of Helsinki*. Full consent was obtained after objectives, procedures and inherent risks were explained to all participants.

### Participants

Community partner organizations recruited older adults to participate in a 12-week walking group intervention by sending emails or distributing flyers to their eligible members. The inclusion criteria were: 1) being ≥ 65 years old; 2) being available for 15 weeks; 3) not using a wheelchair or a white cane (for a visual deficit); 4) being able to collaborate (clinical judgment); 5) having a self-reported fear of falling or reduced walking mobility and 6) being able to practice adapted physical activity.

Fifty-two participants (mean age: 74±6 years) were recruited to follow the intervention during the summer (n=15) and winter (n=37) seasons and completed the baseline assessment. Among these participants, 5 (10%) dropped out before the start of the intervention due to medical condition deterioration (n=3) or no longer being interested (n=2). This dropout occurred only in the winter cohort. Among the participants who started the intervention (n=47), 41 participants completed the intervention and post-assessment [summer (n=15/15: 100%) and winter (n=26/32: 81%); see Figure 1]. Thus, the study retention rate is 78.8 % (n=41/52).

**Figure 1:**
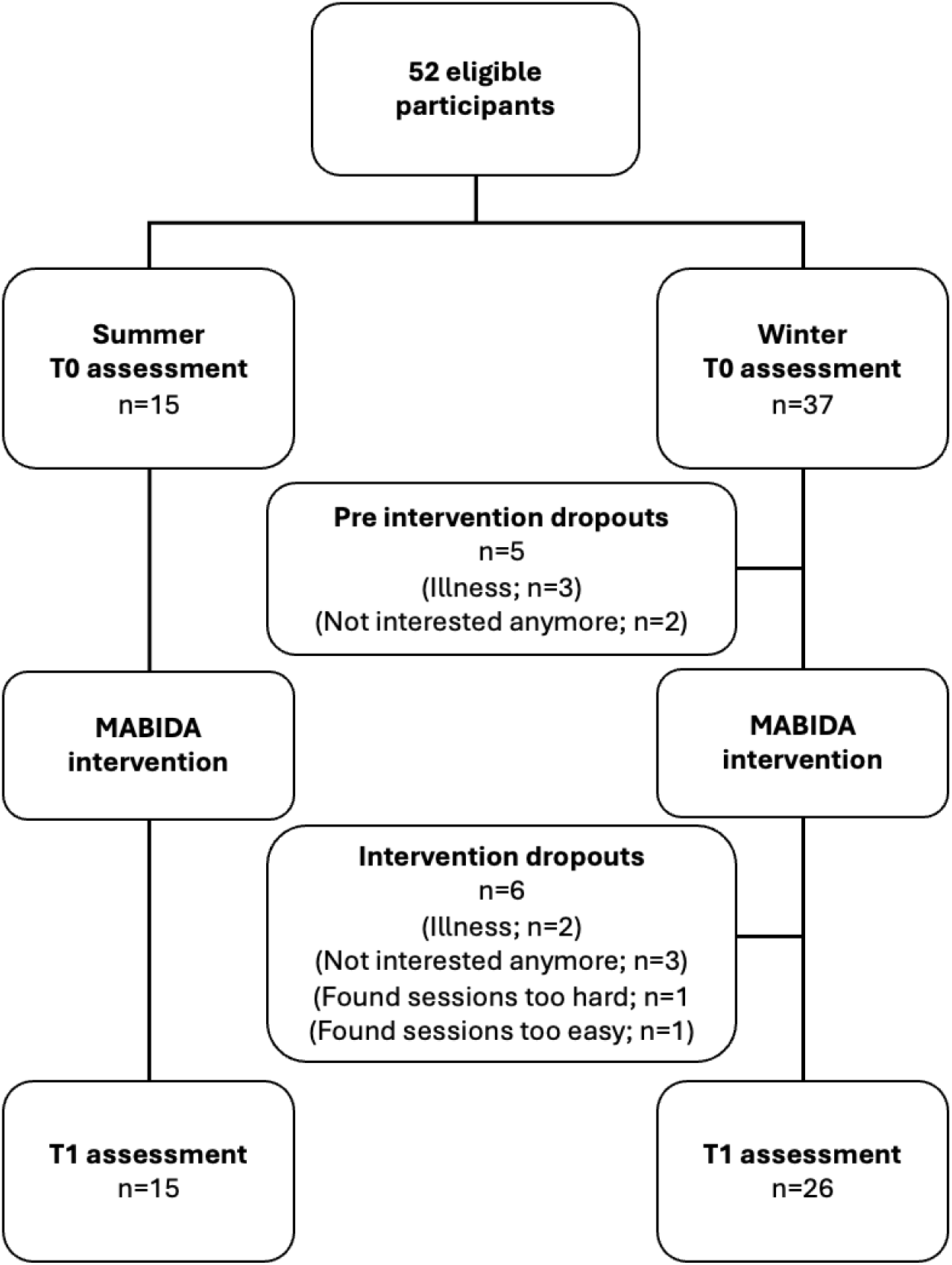
Flow chart of participants.

### MABIDA intervention

#### Step 1: Educational session

As recommended by the literature and the partners, a one-hour group educational session was delivered before the first walking session. The session aimed to increase participants’ awareness of the health benefits of walking and provide practical strategies to facilitate its integration into daily life. It included information on the benefits of walking, proper walking posture, injury prevention, intensity progression, and the use of exercise cards. Participants were also introduced to activity tracking tools to support motivation and informed about community resources available to older adults. Finally, information was presented on public transport services, including free access to public transport for people over 65, and details of training sessions offered by the local public transport provider on how to use public transport safely and confidently. However, regardless of the season, none of the participants chose to take up this free training.

#### Step 2: walking intervention

##### a) Mobility decisional tree

To ensure the appropriateness of the intervention with respect to walking participants’ ability, given that it has been shown to be an implementation barrier among community partners and older adults, a pragmatic decision tree was co-created to evaluate the mobility profile. This decisional tree included 3 questions: Q1 (estimated walking speed): Compared to people my age: I walk as fast or faster (0pt) *or* I walk slower (1pt); Q2 (estimated walking endurance): I walk 10 minutes: with pauses (1pt) *or* without pauses (0pt); Q3 (estimated walking capacity): When I walk fast, I feel out of breath: Yes (1pt) *or* No (0pt). Based on the total score, participants were classified as follows: slow walker (2-3pts) or normal walker (0-1pt). However, to ensure confidence and group integrity/safety, participants had the option to switch groups after the first walk, at their request and per the recommendation of the professional who supervised the walking sessions.

To objectively validate the ability of the mobility decisional tree to classify the participants, they also performed a 3-meter usual pace timed-up and go [TUG (seconds); normal mobility criteria: <10 (Podsiadlo & Richardson, 1991)] and a normal speed 4-meter walking [WS (m/s); normal vs. fast walking speed criteria: 1.00 m/s (Spruijt et al., 2026)] tests. Participants with a TUG <10 sec and WS >1.00 m/s were classified as normal walkers whereas participants with a TUG >10 sec and a WS <0.99 m/s were classified as slow walkers. These validated tests were selected due to their ease of implementation, as they require minimal space and equipment, making them suitable for the future implementation of MABIDA as an appropriate and integrated offer by the partner.

##### b) Walking program

The MABIDA walking program was developed to be given in a wave of 12 consecutive weeks and included one guided walk (1 hour) per week with the participants’ dedicated walking speed group. During this study, given its objective of assessing feasibility, the walks were supervised by a professional (kinesiologist). As mobility in real life setting includes utilitarian and recreational walking, half of the MABIDA walking sessions were conducted in a nearby park, while the other half took place around the community center, using the surrounding streets to access grocery stores and other local amenities. During the walks, the kinesiologist guided participants through various exercises using the city’s furniture (e.g. squats with a bench, balance with something to hold on to if necessary) every 15 minutes for 5 minutes. The distance was increased each week according to the group’s capacity (see Table S-1 in supplemental file for more details). It was also planned to conduct walks and exercises indoors when the weather conditions were considered extreme to ensure safety (temperature too hot or too cold; air pollution). All the walking routes were validated by partners according to participants’ desires. Moreover, in between supervised walks, participants were encouraged to perform at least 40 minutes of walking at moderate intensity (not able to speak continuously) by themselves and to record it in a notebook (duration, distance or step count).

##### c) Exercise cards in-between guided walking sessions

In addition to the unsupervised walks, the participants were encouraged to perform at least 40 minutes of exercise cards per week (free to choose amongst all the cards) and to record it in a notebook. To perform these cards safely and by themselves, participants were trained to perform 6 cards of each type of exercise and intensity (from easy to advanced) during the first 3 guided walks (see Figure S-1 in supplemental file for some examples).

More specifically, these co-created cards (see Figure S-1 in supplemental file) included two modalities (alone (n=66) or in pairs (n=33)), three intensity levels (advanced, intermediate or easy) and three types of exercises [balance (Solo: n=27; Duo: n=12), strength (Solo: n=18; Duo: n=9) or endurance (Solo: n=21; Duo: n=12)]. These cards aimed at improving or avoiding decline in fall predictors such as strength, balance, flexibility and mobility.

### Measures

#### Characterization

Age, sex, body mass index (BMI: kg/m^2^), cognitive status (MoCA: x/30), depression status (GDS-30: x/30) as well as fear of falling (FES-I: x/28 (Kempen et al., 2008)), physical activity status (RAPA: x/10 (Topolski et al., 2006)), life-space mobility (LSA: x/120 (Baker et al., 2003)), functional capacities (Short Physical Battery (SPPB): x/12; pre-disabled: 6<X<10) and gait parameters (3-m TUG: sec (Podsiadlo & Richardson, 1991)) were collected.

#### Implementation variables

##### - Feasibility variables

Retention rate was calculated for the whole cohort to measure the number of participants who completed the study divided by the number of participants recruited.

Adherence to the supervised intervention portion was measured throughout the study via the ratio of presence to supervised weekly walks (%: n/12 weeks). A ratio ≥ 75% was set as acceptable.

Adherence to the self-reported supervised intervention portion was assessed using the notebook and estimated with the completion of challenge cards (%: x/40 minutes per week/12 weeks) and the time of walk (%: x/40 minutes per week/12 weeks) every week in between the supervised sessions. Being non-supervised and voluntary, the percentage of exercises performed outside the group sessions was considered acceptable if x≥ 50%.

Feasibility of the program was evaluated through 4 items using a Likert scale every three weeks. Specifically, participants rated their agreement (1: completely disagree, 2: disagree, 3: agree, 4: completely agree) to the following statements: 1) *difficulty*: The level of difficulty of the group walk suits me; 2) *hour*: The hour of the session suits me; 3) *day*: The day of the session suits me; and, 4) *duration*: The duration of the session suits me. A score x>75% was considered very feasible, a score 50% >x< 75% was considered feasible, and x< 50% was considered non-suitable.

#### Acceptability variables

Participants’ acceptability was recorded every 3-weeks through 4 items (satisfaction, enjoyment, wellbeing, delivery appreciation) using a Likert scale (1: completely disagree, 2: disagree, 3: agree, 4: completely agree). Statements were as follows: satisfaction «I am satisfied with the session»; enjoyment «I enjoyed the session»; wellbeing «I felt good after participating in the session»; delivery «I liked the way the session was conducted». A score x>75% was considered very acceptable, a score 50% >x< 75% was considered acceptable, and x< 50% was considered non-acceptable.

Barriers and facilitators were determined with open questions at the last assessment and participants’ written commentaries in their notebook.

### Data analysis and statistics

Due to our objectives and design (pilot study), a per-protocol analysis was performed with participants who completed the final assessment (independently of adherence). Descriptive statistics were used to characterize each cohort (summer and winter) and each group (normal and slow). To account for the small sample size (phase II pilot study) and possible abnormal distribution, quantitative data were compared with a Mann-Whitney test (Winter vs. Summer; Slow vs. Normal; ALL completed vs. dropouts). A Fisher’s exact probability test was used to evaluate the categorical data regarding feasibility and acceptability. P <0.05 was considered significant (IBM SPSS Statistics 31.0 (SPSS Inc., Chicago, IL)).

## RESULTS

### Baseline characteristics

MABIDA participants had a mean age of 74.0±5.8 years, and 17% were men. Age, BMI, cognition, depression status, physical activity level, life-space assessment, and fear of falling were similar between seasonal and walking groups (p>0.05; see Table 1). At baseline, SPPB score was 9.9±1.4 for the entire cohort, with no significant difference (p>0.05) between seasons. Walking speed only was statistically different between summer (n=15) and winter (n=26) groups (p<0.05). Per design, usual 3m-TUG, 5-STS, walking speed and SPPB were lower in the slow walking group (SWG: n=19) than in the normal walking group (NWG: n=22; p<0.05; effect size >0.3<0.5; see Table 1). Finally, baseline characteristics were also similar (p>0.05) between those who completed the study (n=41) and those who dropped out (n=11; see Table 1).

**Table 1:**
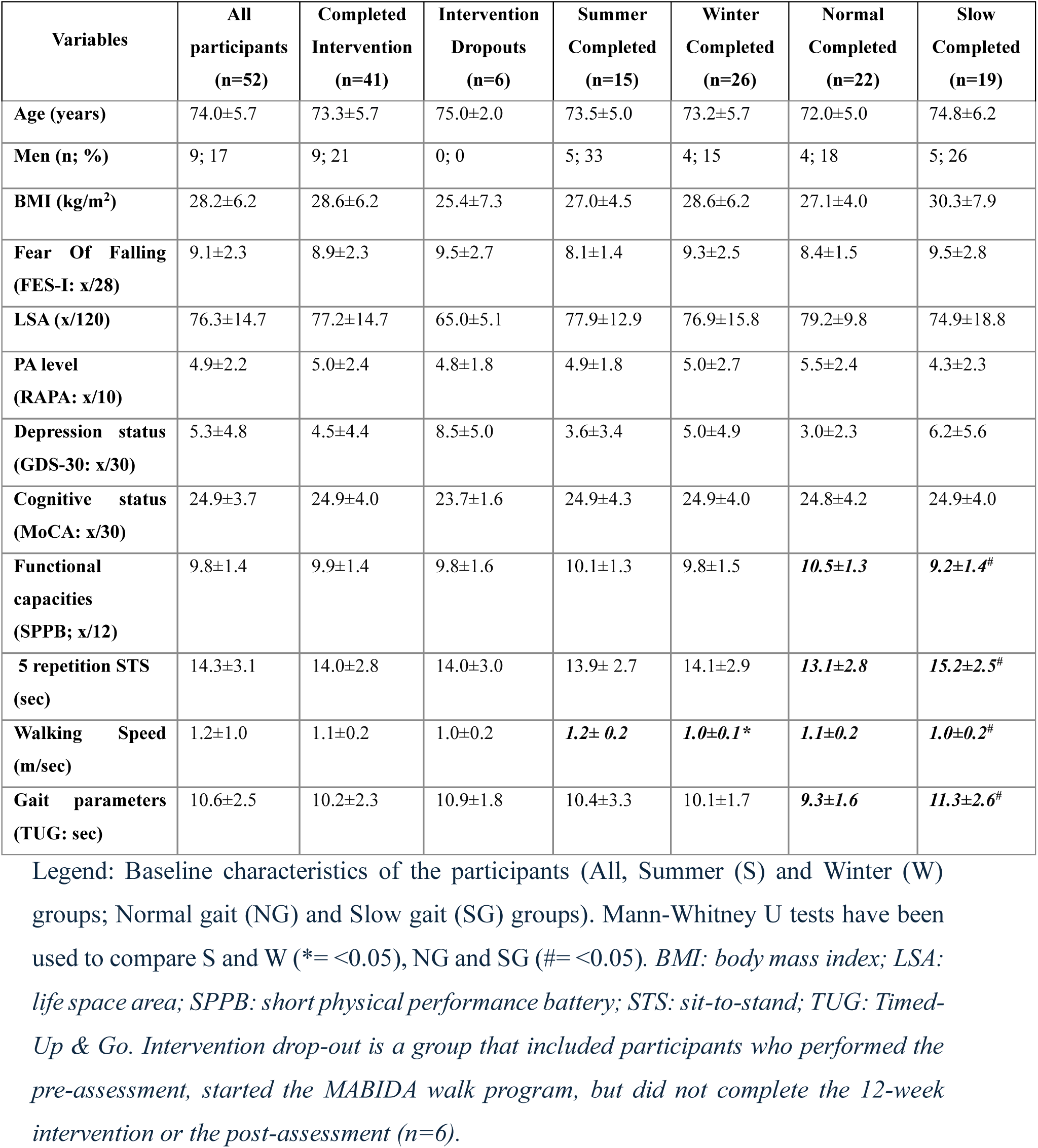
Baseline characteristics of the participants.

### Feasibility

Regarding the mobility decisional tree, among the participants who started the intervention (n=47), 70% of those initially classified in the normal walking group (n=23/33), and 79% of those classified in the slow walking group (n=11/14) were confirmed in their respective walking group after the objective assessments (see supplemental Figure S-2). Thus, the pragmatic decision tree correctly classified 34 of 47 participants (72%) compared with the objective assessments. No difference in attrition during the intervention was observed between those well classified (NWG: 83%; SWG:82%) and those not well classified (NWG or SWG: 100%).

Regarding the retention rate, 78.8% of the participants recruited completed the study (n=41/52), and 87% of those who started the intervention (n=41/47) completed the study. All the dropouts during the intervention (n=6; NWG: n=4 vs. SWG: n=2) occurred during the winter cohort [not interested anymore: n=3 (NWG), illness: n=1 (SWG), found the sessions too easy: n=1 (NWG), found the sessions too hard: n=1(SWG)].

Regarding supervised intervention adherence, the whole group completed 86±12% of the sessions with no difference between the seasons (summer: 87.0±8.5% vs. winter: 85.3 ± 13.0%; p=0.90; see supplementals for weather adherence) or between the walking speed groups (NWG: 88.0±7.4% vs SWG: 84.0±14.8%, p=0.55).

For self-reported adherence, 87% of all participants completed, on average, more than 40 min/week of walking over the 12-week intervention. However, only 24% of them completed, on average, more than 40 min/week of challenge card exercises over the 12-week intervention. No difference between the seasonal (walking time: summer:93% vs. winter:85%; cards: summer:27% vs. winter:23%) or between the walking speed (walking time: NWG:90% vs. SWG:84%; cards: NWG:32% vs. SWG:16%) groups (p>0.05).

Considering the intervention feasibility (see Table 2), the frequency (day), time (hour) and duration were found to be appropriate by 91%, 82% and 81% of all participants, respectively. Overall, 84% agreed or completely agreed that the difficulty of the walking sessions suited them. There was no statistical difference between summer and winter groups (p>0.5) nor between slow and normal walking groups (p>0.05).

**Table 2:** Acceptability and feasibility of the MABIDA intervention for each group according to participants’ perceptions.

| ACCEPTABILITY |  |  |  |  |  |
| --- | --- | --- | --- | --- | --- |
|  | All | Summer | Winter | Normal | Slow |
| **Satisfaction** |  |  |  |  |  |
| Completely agree (%) | 66 | 68 | 63 | 73 | 59 |
| Agree (%) | 16 | 23 | 12 | 18 | 13 |
| Disagree (%) | 2 | 0 | 3 | 2 | 1 |
| Completely disagree (%) | 0 | 0 | 0 | 0 | 0 |
| Did not respond (%) | 16 | 9 | 22 | 7 | 27 |
| **Enjoyment** |  |  |  |  |  |
| Completely agree (%) | 64 | 83 | 53 | 76 | 50 |
| Agree (%) | 17 | 15 | 18 | 15 | 20 |
| Disagree (%) | 3 | 0 | 5 | 3 | 3 |
| Completely disagree (%) | 1 | 0 | 1 | 0 | 1 |
| Did not respond (%) | 14 | 2 | 23 | 6 | 26 |
| **Wellbeing** |  |  |  |  |  |
| Completely agree (%) | 70 | 87 | 60 | 82 | 55 |
| Agree (%) | 13 | 12 | 14 | 13 | 14 |
| Disagree (%) | 2 | 0 | 3 | 0 | 4 |
| Completely disagree (%) | 0 | 0 | 0 | 0 | 0 |
| Did not respond (%) | 15 | 1 | 23 | 5 | 27 |
| **Delivery** |  |  |  |  |  |
| Completely agree (%) | 66 | 75 | 61 | 73 | 58 |
| Agree (%) | 17 | 23 | 13 | 20 | 13 |
| Disagree (%) | 2 | 0 | 3 | 1 | 3 |
| Completely disagree (%) | 0 | 0 | 0 | 0 | 0 |
| Did not respond (%) | 15 | 2 | 23 | 6 | 26 |

FEASIBILITY
|  | All | Summer | Winter | Normal | Slow |
| --- | --- | --- | --- | --- | --- |
| **Difficulty** |  |  |  |  |  |
| *Completely agree (%)* | 55 | 60 | 52 | 61 | 47 |
| *Agree (%)* | 29 | 38 | 23 | 33 | 24 |
| *Disagree (%)* | 1 | 0 | 1 | 0 | 1 |
| *Completely disagree (%)* | 1 | 0 | 1 | 0 | 1 |
| *Did not respond (%)* | 14 | 2 | 23 | 6 | 27 |
| **Day** |  |  |  |  |  |
| *Completely agree (%)* | 75 | 83 | 70 | 82 | 67 |
| *Agree (%)* | 9 | 12 | 7 | 10 | 7 |
| *Disagree (%)* | 0 | 0 | 0 | 0 | 0 |
| *Completely disagree (%)* | 0 | 0 | 0 | 0 | 0 |
| *Did not respond (%)* | 16 | 5 | 23 | 8 | 26 |
| **Hour** |  |  |  |  |  |
| *Completely agree (%)* | 73 | 85 | 65 | 84 | 59 |
| *Agree (%)* | 9 | 13 | 7 | 10 | 8 |
| *Disagree (%)* | 3 | 0 | 5 | 0 | 7 |
| *Completely disagree (%)* | 0 | 0 | 0 | 0 | 0 |
| *Did not respond (%)* | **15** | **2** | **23** | **6** | **26** |
| **Duration** |  |  |  |  |  |
| *Completely agree (%)* | **65** | **73** | **61** | **76** | **53** |
| *Agree (%)* | **16** | **20** | **14** | **17** | **16** |
| *Disagree (%)* | **2** | **2** | **2** | **1** | **3** |
| *Completely disagree (%)* | **0** | **0** | **0** | **0** | **0** |
| *Did not respond (%)* | **17** | **5** | **23** | **9** | **28** |

*Legend: Responses are presented as percentages for each response category from the Likert scales. “Normal” and “Slow” refer to participants’ walking-speed groups, while “Summer” and “Winter” refer to the intervention season. “Did not respond” indicates missing responses to the corresponding item.*

### Acceptability

Overall, 82% of the participants found the intervention satisfying/very satisfying and 81% enjoyable/very enjoyable (see table 2). Eighty-three percent of them found that they agreed or completely agreed that they felt good after the walking sessions, and 83% found that they liked the way the sessions were conducted. No seasonal or walking groups differences (p>0.05) were observed [1) very satisfied/satisfied: winter:75% *vs.* summer:91% or SWG:72% *vs.* NWG:91%; 2) enjoyable/very enjoyable: winter:71% *vs.* summer:98% or SWG:70% *vs.* NWG:91%; 3) feeling good/very good: winter:74% *vs.* summer:99% or SWG:69% *vs.* NWG:95%; 4) appreciated a lot: winter:74% *vs.* summer:98% or SWG:71% *vs.* NWG:93%]. One person (from the winter/slow walking group) did not respond.

#### Barriers and facilitators (see Figure 4)

**Figure 4-A:**
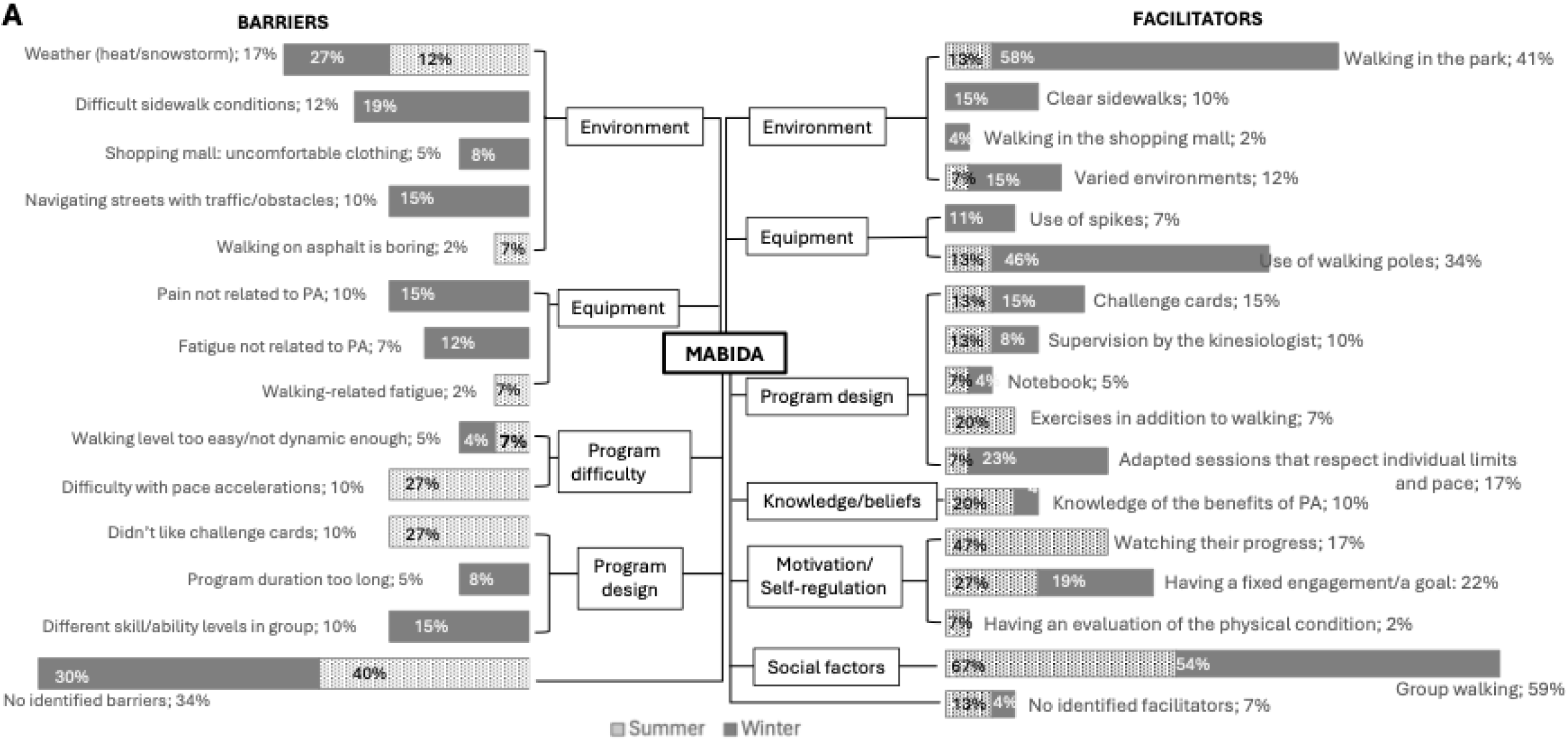
Barriers and facilitators related to MABIDA implementation for Summer and Winter groups. Legend: The percentage is presented for the whole group in the label, and for each group (Summer-Winter) in their bars

**Figure 4-B:**
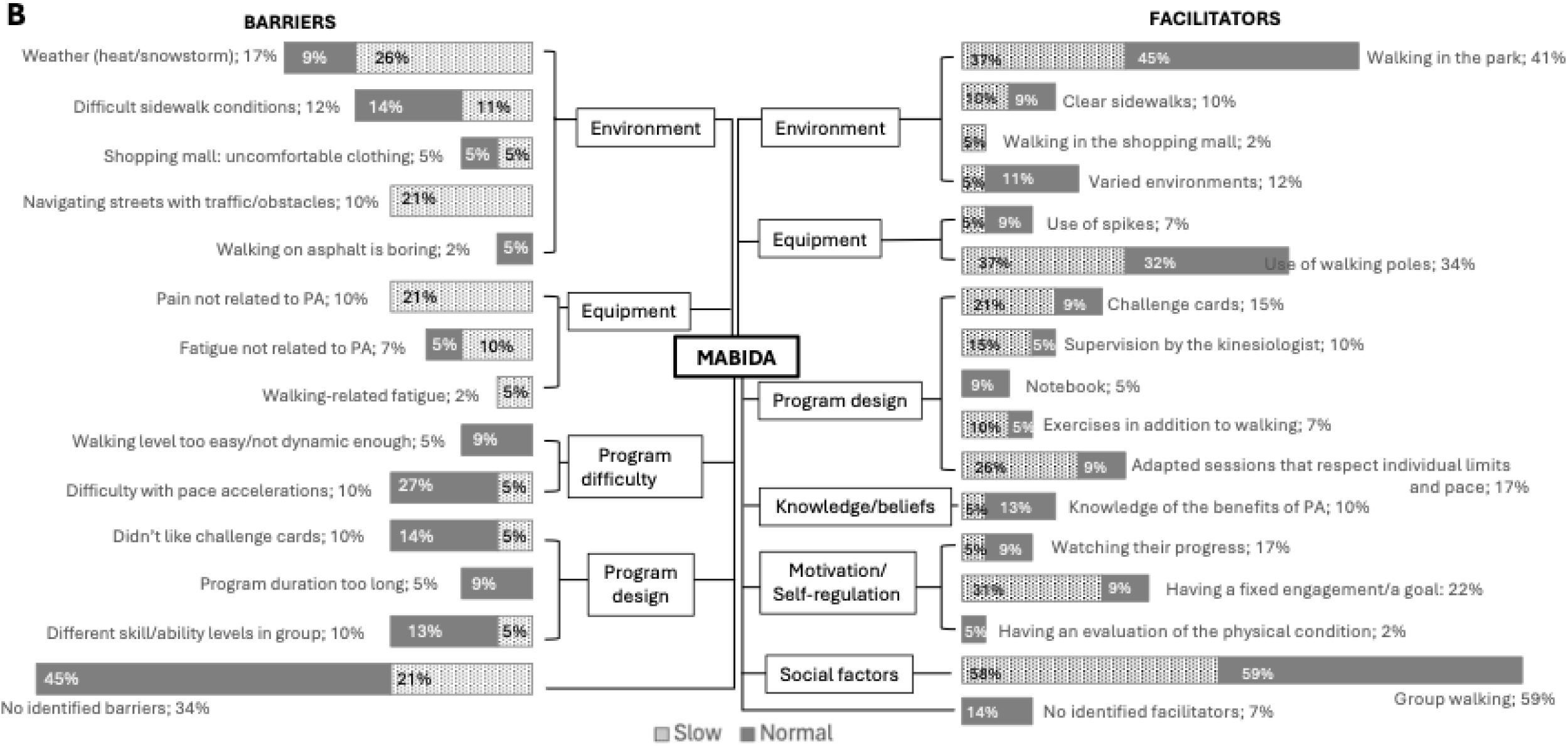
Barriers and facilitators related to MABIDA implementation for Normal & Slow walking groups. Legend: The percentage is presented for the whole group in the label, and for each group (Normal-Slow) in their bars.

Among the participants, 34% didn’t identify any barriers to the program. The main barrier was the weather (overall:17%). More specifically, 40% of the summer participants were mainly concerned about heat and humidity, and 36% of the winter participants were concerned about snow and ice. The other barriers were related to the season. During winter, barriers were mostly environmental [navigating streets with traffic and obstacles (15%); difficult sidewalk conditions (19%)] and physical [fatigue (12%) and pain (15%) not related to PA] whereas during summer, the most-mentioned barriers were in relation to the program design [didn’t like challenge cards (27%); difficulty with pace accelerations (27%)]. For the walking groups, temperature was the highest barrier in the SWG (26% vs. NWG:9%), followed by navigating streets with traffic and obstacles (21% vs. NWG:0%) and pain not related to PA (21% vs. NWG:0%). Forty-five percent of the normal gait group reported no barriers, compared with 21% in the slow walking group.

Regarding facilitators, the main one identified by participants overall (59%), independently of season (summer:67% vs. winter:54%) or walking group (SWG:58% vs. NWG:59%), was social (walking in a group). Having a fixed engagement or a goal, as well as walking in the park, were also important for all the participants (22% & 42% respectively) independently of the season (summer:27% vs. winter:19% & summer:13% vs. winter:58% respectively) or the walking group (SWG:39% vs. NWG:45%).

## DISCUSSION

The main objectives of this project were to assess the feasibility and acceptability of implementing the community setting MABIDA walking program in older adults with self-reported fear of falling.

First, regarding the implementation of MABIDA, its rate was comparable to or higher than those reported for similar interventions or comparable populations (Weber et al., 2018), although all dropouts after the start of the intervention occurred during the winter (Picorelli et al., 2014). More specifically, most of these dropouts were in the normal walking group (n=4/6), which is contrary to other studies that commonly reported this pattern in participants with poorer health or lower physical function (Picorelli et al., 2014). The reasons provided (“too easy” and “loss of interest”) suggest that withdrawal might have been driven by insufficient challenge or declining motivation. This interpretation is consistent with previous evidence indicating that interventions perceived as enjoyable, beneficial for them, sufficiently challenging and adapted to the participant’s abilities have a better attrition rate (Farrance et al., 2016; Picorelli et al., 2014; Yang et al., 2024). As reported by Delaire et al. (Delaire et al., 2021), another potential explanation is that older adults with poorer physical function may have perceived greater health benefits from the intervention, whereas those with fewer deficits may not have perceived it as necessary for maintaining or improving their quality of life.

Adherence to the supervised walking sessions was high across all groups, and a large proportion of participants achieved the voluntary target of an average of 40 minutes of walking per week over the 12-week intervention. Together, these findings suggest that the walking component of the program was feasible and well accepted by participants. However, only 24% of the participants completed the recommended amount of challenge card exercises. This lower adherence could be partly explained by the fact that many participants reported engaging in other forms of exercise such as physiotherapy, yoga, or dance instead of the challenge card exercises. It may also reflect aspects of the implementation itself, as the professional delivering the intervention and the educational session may not have explained the role or importance of these cards for preventing fall-related risk factors enough. Thus, before implementing MABIDA at a larger scale, it will be important to determine whether modifications to the exercise cards’ content, format, or integration into the intervention could improve adherence. These findings also highlight the importance of considering the feasibility of individual intervention components rather than the program itself. While adherence to the walking components was high, the exercise-card component appears to require further refinement before larger-scale implementation.

Furthermore, the intervention settings (frequency (day), time (hour) and duration) were considered very appropriate for the participants, who found that the difficulty was well adapted to their abilities, independently of the sub-groups. These results suggest that the intervention is feasible and well adapted for older adults living in the community with a fear of falling. These findings are important and relevant, as appropriate exercise intensity and convenience have been identified in systematic reviews of community-based exercise interventions for older adults as predictors of adherence (Farrance et al., 2016; Picorelli et al., 2014; Yang et al., 2024).

Moreover, to ensure safety, suitability and inclusivity for all participants, it is important to have specific group sessions. The implementation of a simple three-question subjective decision tree showed promising potential as a pragmatic tool for assigning participants to walking groups, with 72% (n=34/47) of participants classified consistently with the objective assessments. Given its simplicity and ease of administration, this tool may facilitate group allocation in community settings. However, further validation is needed, particularly given the potential implications of misclassification for participant safety and appropriate intervention tailoring.

Participants of the MABIDA program also had a high level of satisfaction and enjoyment, and they experienced a sense of wellbeing after the sessions. These results are important as these aspects contributed to physical activity participation and motivation (Devereux-Fitzgerald et al., 2016). Delivery was also well received, as 83% liked the way the sessions were conducted. Moreover, most participants reported appreciating the listening skills and adaptability of the kinesiologist, which made them feel confident in their abilities, rendering the activity accessible. This result aligns with evidence showing that instructor behaviour contributes to adherence (Farrance et al., 2016; Yang et al., 2024).

Regarding the modality of the program proposed, the main facilitator was the group setting (58%). This finding is in accordance with studies showing that social factors can be a determinant in motivation (Devereux-Fitzgerald et al., 2016). However, it is important to mention that the nature of the intervention is likely to attract individuals who are more prone to liking social settings and to be looking for group activities. Furthermore, half of the intervention was carried out in the park (as opposed to walking in the city streets), which was the second motivator. This aspect is important as it has been shown that exposure to greenery and nature positively influences PA and walking in older adults (Stearns et al., 2023). Furthermore, the teaching and use of equipment (e.g. walking poles) were also another relevant aspect of the MABIDA intervention, as it has been reported that it might reduce fear of falling, as it is an aid to walking and increases stability or balance during walking (Bateni & Maki, 2005) or to negotiate environmental obstacles like iced sidewalks, uneven surfaces and snow accumulation (Li et al., 2013).

Regarding its implementation in a northern country, although winter is generally associated with lower PA and mobility levels in older adults (Jones et al., 2017; Portegijs et al., 2014), winter weather didn’t seem to be generally perceived as a barrier by the participants. In fact, weather was mentioned more often in the summer group (heat and humidity; 27%) than in the winter group (snow and ice; 12%) as a barrier. This is consistent with studies showing that heat and humidity have an impact on physiology (Meade et al., 2020) and can lower PA levels in older adults (Jones et al., 2017). One hypothesis for the seasonal difference could be that it is easier to adjust clothing in winter (adding layers, wearing appropriate materials) than in summer to be comfortable for practicing PA.

Besides weather and «walking level too easy», the barriers mentioned differed from summer to winter, suggesting that barriers must be approached differently depending on the season. For example, conditions of the pedestrian facilities have been reported in the literature to be a barrier to walking mobility in winter for older adults (Bergen et al., 2023; Clarke et al., 2017; Li et al., 2013; Wennberg et al., 2009). However, our results suggest that sidewalk conditions might indeed be a greater deterrent to walking (19%) than winter weather (12%). Although these factors are related—since weather affects sidewalk conditions—the latter can be mitigated through municipal action. Consistent with prior findings that slippery or difficult surfaces pose a greater barrier than temperature alone (Li et al., 2013), our results suggest that 1) planning walking routes in areas where municipal maintenance is faster and 2) collaborating with the municipality to prioritize prompt sidewalk maintenance would support more successful implementation during winter.

This study presents some limitations. First, as a pilot study, MABIDA was primarily designed to assess the feasibility and acceptability of the intervention. Thus, the use of a convenience sample and the relatively small sample size limit the generalizability of the findings to the broader population of community-dwelling older adults and did not allow us to evaluate the season*walking group interaction. In addition, the intervention was implemented in a single municipality, which may further limit the transferability of the results to other geographical or organizational contexts. For safety reasons due to the research stage, the intervention was provided by an external and trained resource. Nevertheless, one of MABIDA’s strengths is its development using a co-designed approach involving key stakeholders, allowing the intervention to be informed by the needs, preferences and capacities of older adults as well as the practical considerations relevant to community-based implementation. Furthermore, the pragmatic nature of the intervention, delivered under real-world conditions (step 1 of Action-oriented research), supports its potential for future implementation in real-life settings. Nevertheless, by design, real-life scale-up research fully led by the community partner (e.g. educational sessions; group assignment; walking sessions guided by a peer; intervention modality, etc.) is needed to confirm its implementation and to produce a knowledge transfer guide for community partners.

## CONCLUSION

Overall, the findings support that the MABIDA walkability program seems feasible, acceptable and appropriate for older adults living in the community and reporting being afraid of falling, independently of the season or the walking speed profile. These promising results support further collaboration with local partners (city and organizations) to refine the program and strengthen its sustainability and alignment with community implementation needs and resources. Future research should also examine the potential effects of MABIDA on fall-related predictors (i.e., dose-response) and assess its feasibility and sustainability when implemented on a real-life scale-up.

## Acknowledgments

We would like to express our gratitude to the participants, the city of Longueuil, the community organization, the kinesiologists and the research assistants who participated in this study and made it possible to explore the integration of this intervention into a real-life setting.

## Author’s contribution

ML (1^st^ author): performed the data curation, performed the statistical analyses, wrote and revised the manuscript. MAL (senior author): Conceptualization, Tool co-creation, Methodology, Investigation, Visualization, Writing - Review & Editing, Validation, and Supervision. ML, FR & MSC: conceptualized the main study and reviewed the manuscript. KL: participated as a partner and reviewed the manuscript. HB: conducted data collection, helped with data curation and reviewed the manuscript.

## Data availability

The datasets generated during and/or analyzed during the current study are available from the corresponding author on request.

## Statement of AI use

AI tools were used exclusively for English language editing. AI tools were not used for data analysis, interpretation, or the generation of scientific writing content.

