## Supplementals for "Feasibility & Acceptability of a co-created pragmatic walkability program to promote mobility in older adults with fear of falling: *A question of season and mobility profile?*"

### SUPPLEMENTAL MATERIALS

#### Table S-1: MABIDA walking program implementation

| *Week Session* | *Warm-up* | *Walking program* | *Cool-down* |
|---|---|---|---|
| ***Weeks 1–3*** | 5 min | 1) Comfortable and continuous walk: 10 min**<br>2) Exercise cards (3 cards): 10 min#<br>3) Comfortable and continuous walk: 10 min**<br>4) Exercise cards (3 other cards):10 min#<br>5) Comfortable and continuous walk :10 min**<br>***Borg RPE: week 1: 9-10/20; week 2: 10-11/20; week 3: 11-12/20*<br>*# Cards: week 1: balance exercise; week 2: strength exercise; week 3: endurance exercise* | 5 min |
| ***Weeks 4–6*** | 4 min | 1) HIIT walk (Borg RPE: 13-14/20): 16 min**<br>2) Comfortable and continuous walk (RPE 10-11) : 10 min<br>3) HIIT walk (Borg RPE: 13-14/20): 16 min**<br>4) Comfortable walk (RPE 10-11): 10 min<br><br>**HIIT details (2 min/ cycle* 8 cycles):<br>week 4: fast (Borg RPE: 14-16/20): 30 sec + normal (Borg RPE: 12-13/20): 90 sec;<br>week 5: fast (Borg RPE: 14-16/20): 60 sec + normal (Borg RPE: 12-13/20):60 sec;<br>week 6: fast (Borg RPE: 14-16/20): 90 sec + normal (Borg RPE: 12-13/20):30 sec. | 4 min |
| ***Weeks 7–9*** | 5 min | 1) Comfortable and continuous walk with poles:10 min **<br>2) Exercise with poles: 10 min<br>3) Comfortable and continuous walk with poles: 10 min **<br>4) Exercise with poles: 10 min<br>5) comfortable and continuous walk with poles: 10 min**<br><br>**Borg RPE: week 7: 10-11/20; week 8: 11-12/10; week 9: 12-13/20 | 5 min |

| | | | |
|---|---|---|---|
| ***Weeks 10–12*** | 5 min | Brisk walk: 50 min ** <br> **Borg RPE: week 10: 13-14/20; week 11: 14-15/20; week 3: 15-16/20 | 5 min |

<u>*Legend:*</u> instructions for Borg RPE from 9 to 12/20: Ensure that you can speak easily while walking (light intensity); instructions for borg from 12 to 14: Increase the pace so you feel short of breath but still able to speak a full sentence (moderate intensity). Instructions for borg from 14 to 16: Increase the pace until you feel difficulty speaking a full sentence (moderate to high intensity). Week 4: Familiarization with the interval walking cycles. Week 7: Familiarization with the use, posture, and adjustment of the walking poles. Note: During winter, walking with poles started from week 1. The description included park and city walk implementation.

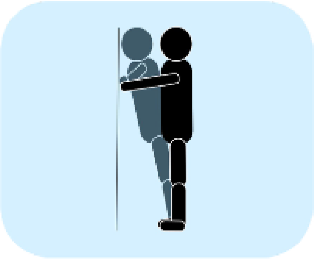

**Wall push-ups**

Lower your body toward the wall by bending your elbows and keeping your back straight.
Push against the wall to return to the starting position.

→ Repeat 10 times

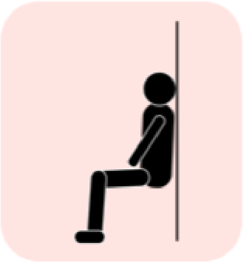

**Chair on wall**

Bend your knees until your thighs are parallel to the floor and hold the position while keeping your back straight.

→ Stay 30 seconds

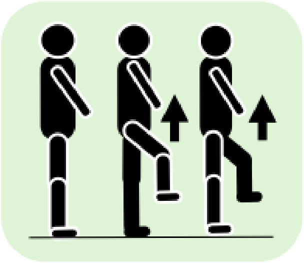

**Walk with knee flexion**

Move forward while keeping your back straight and bending each knee to 90 degrees with every step.

→ Repeat 10 times on each side, alternating sides

*Figure S-1: Example of solo challenge cards*

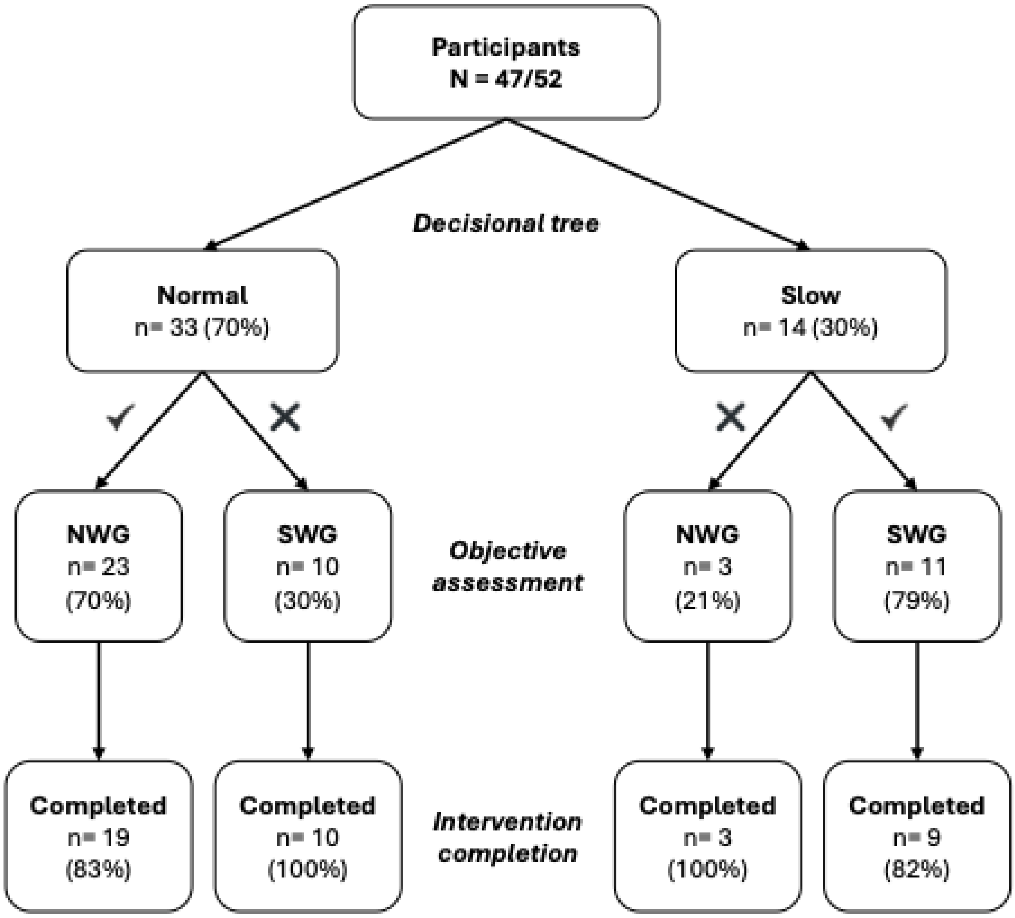


**Figure S-2 Decisional tree assignment flow chart**

*Legend: NWG: participants included in normal walking group; SWG: participants included in slow walking group.*

**Table S-2A: Weather and adherence for the summer group**

| Summer | | | | |
|---|---|---|---|---|
| | **Presence (%)** | **Temperature (°C)** | **Weather** | **Walking location** |
| **Week 1** | **93** | **20** | **Clear** | **City** |
| **Week 2** | **93** | **25** | **Clear** | **Park** |
| **Week 3** | **100** | **20** | **Clear** | **City** |
| **Week 4** | **80** | **30** | **Heatwave** | **Park** |
| **Week 5** | **93** | **26** | **Clear** | **City** |
| **Week 6** | **87** | **27** | **Clear** | **Park** |
| **Week 7** | **93** | **28** | **Clear** | **City** |
| **Week 8** | **80** | **26** | **Clear** | **Park** |
| **Week 9** | **93** | **22** | **Clear** | **City** |
| **Week 10** | **93** | **27** | **Humidity** | **Park** |
| **Week 11** | **93** | **22** | **Clear** | **City** |
| **Week 12** | **67** | **23** | **Inundations (154mm)** | **Park** |

*Legend: % of participants present at the group walking session, temperature, weather and walking location for each week of the summer intervention. Temperature was collected by the instructor during the session through the Atmotube pro (Atmotube©).*

**Table S-2B: Weather and adherence for the winter group**

| Winter | | | | |
|---|---|---|---|---|
| | **Presence (%)** | **Temperature (°C)** | **Weather** | **Walking location** |
| **Week 1** | **92** | **-10** | **Clear** | **city** |
| **Week 2** | **85** | **-16** | **Clear** | **City** |
| **Week 3** | **92** | **-15** | **Ice** | **Park** |
| **Week 4** | **89** | **-16** | **Clear** | **Park** |
| **Week 5** | **89** | **-12** | **Snowstorm (34 cm of snow)** | **City** |
| **Week 6** | **69** | **-14** | **Snowstorm (36 cm of snow)** | **Park** |
| **Week 7** | **89** | **-4** | **Snow** | **Park** |
| **Week 8** | **96** | **0** | **Rain** | **Shopping mall** |
| **Week 9** | **77** | **-10** | **Ice** | **City** |
| **Week 10** | **89** | **-3** | **Clear** | **City** |
| **Week 11** | **85** | **1** | **Clear** | **Park** |
| **Week 12** | **73** | **-3** | **clear** | **City** |

*Legend: % of participants present at the group walking session, temperature, weather and walking location for each week of the winter intervention. Temperature was collected by the instructor during the session through the Atmotube pro (Atmotube©).*
